# Geographic Barriers to Achieving Universal Health Coverage in a rural district of Madagascar

**DOI:** 10.1101/2020.07.15.20155002

**Authors:** Andres Garchitorena, Felana A Ihantamalala, Christophe Révillion, Laura F Cordier, Mauricianot Randriamihaja, Benedicte Razafinjato, Feno H Rafenoarivamalala, Karen E. Finnegan, Jean-Claude Andrianirinarison, Julio Rakotonirina, Vincent Herbreteau, Matthew H Bonds

**Affiliations:** MIVEGEC, Univ. Montpellier, CNRS, IRD, Montpellier, France; NGO PIVOT, Ranomafana, Madagascar; Université de La Réunion, UMR 228 Espace-Dev (IRD, UA, UG, UM, UR), Saint-Pierre, Réunion, France; School of Management and Technological innovation, University of Fianarantsoa, Madagascar; Department of Global Health and Social Medicine, Harvard Medical School, Boston, USA; Ministry of Public Health, Antananarivo, Madagascar; National Institut of Public Health, Antananarivo, Madagascar; Faculty of Medicine, Antananarivo, Madagascar; Institut de Recherche pour le Développement, UMR 228 Espace-Dev (IRD, UA, UG, UM, UR), Phnom Penh, Cambodia

## Abstract

Poor geographic access can persist even when affordable and well-functioning health systems are in place, limiting efforts for universal health coverage (UHC). It is unclear how health facilities and community health workers contribute to achieving UHC. Using geographic information from thousands of patients in a rural district of Madagascar we evaluate how a health system strengthening (HSS) intervention aimed towards UHC affects the geography of primary care access. We find that facility-based interventions (user-fee exemptions, improved readiness) achieved high utilization rates only among populations who lived in close proximity to supported facilities. Scaling only facility-based HSS programs across the district would result in large gaps in health care access for the majority of the population. Community health provided major improvements in service utilization for children regardless of their distance from facilities. Our results have implications for UHC policies and suggest that greater emphasis on professionalized community health programs is essential.

## INTRODUCTION

Despite considerable progress on the health-related development goals, every year five million children under five die of treatable illness such as malaria, diarrhoea, and respiratory infections. More than three and a half billion people lack access to essential health services [1,2]. At the recent 40-year anniversary of the Alma Ata Declaration, 134 countries signed on to a renewed commitment to Universal Health Coverage (UHC) based on a shared vision that primary health care and health services be “high quality, safe, comprehensive, integrated, accessible, available, and affordable for everyone everywhere” [3]. In practice, UHC policies tend to focus on financial coverage, such as through health insurance, which reduce point-of-service payments that are known to be barriers to care [4–10]. However, there is growing recognition that among the greatest challenges to accessing health care are geographic barriers: terrain, waterways, and other factors associated with physical distance between the patient and the service [11–15]. The use of primary care decreases exponentially for populations living at increasing distance of primary healthcare centers (PHC), known as the “distance decay” effect [11–15]. Distance decay in health access is equivalent to the effect of user fees [16], which can be more directly reduced or eliminated [4–10].

While there is a considerable body of evidence on the relationship between health system access and user fees, there is surprisingly little evidence on the relationship between health system change and the geography of health access. Studies suggest that geographic barriers to PHC persist even when user fees have been removed, making these approaches insufficient to reach full population coverage of primary care services [17–19]. The leading policy strategy for addressing geographic barriers is through community health workers; i.e., lay people who are trained to treat a subset of clinical cases [20,21]. Yet, little is known about the effects of community health (CH) systems on the geography of health access. Can the leading policies designed to improve health care coverage - UHC and CH – actually overcome these key barriers?

Here, we take advantage of a natural experiment in health system strengthening (HSS) in a rural district of Madagascar, a country with one of the least-funded health systems in the world [22] and where the majority of the population live more than 5km from a health facility [5,23,24]. Starting in 2014, the nongovernmental organization, PIVOT, partnered with the government of Madagascar to establish a model health system in the southeastern district of Ifanadiana (∼200,000 people). A range of HSS programs were initiated in a third of the district (see Table S1), ensuring health system readiness (infrastructure, personnel, supply chain, user fee removal), improving clinical programs (maternal and child health) and supporting integrated information systems at all levels of care (community health, primary care facilities, and the district hospital). Early results showed improved quality of primary care [25], a tripling of facility utilization rates [26], and declines in under-five and infant mortality rates in the first two years of intervention [26]. Later, in 2016, these programs were extended to include strengthened community health workers - who were trained, supervised, and equipped [38-43].

With a unique geographically explicit patient-level dataset encompassing all health center visits in the district during four years, we examine how the effects of distance from health facilities on consultation rates change over time, and how these, in turn, are affected by strengthened health programs. The results show that utilization rates decline dramatically within the first 5km even after the system is strengthened at the facility level; those living 5km from a strengthened health facility access care at rates comparable to those living near a facility that did not receive the intervention. We predict that UHC alone would only result in modest increases in primary care coverage. Subsequent support to community health provided major improvements in service utilization for children regardless of their distance from a facility. This is strong evidence that combining UHC policies at health facilities with strengthened community health programs can have substantial impacts on the geographic reach of the health system. More generally, we show how a model system in global health – based on dynamic integration of data from services at multiple levels of the health system – can address fundamental questions on some of the most challenging global health policy issues.

## RESULTS

### PHC utilization by geographic proximity

We obtained information on the Fokontany of residence for all patients attending one of the 19 PHC in Ifanadiana district between January 2014 and December 2017, which we aggregated into monthly per capita visits by Fokontany (Figure 1; see “Methods” section). Of the 314,443 patients who attended a PHC for an outpatient visit, 276,865 patients had a known geographic location and 99.25% of these (274,798) came from within the district (Figure S1). We obtained the shortest path distance from each Fokontany to the nearest health center following a complete participatory mapping of all footpaths (23,726 km), buildings (106,171) and residential areas (4,925) on OpenStreetMap, and calibrated travel time estimations with remote sensing analyses and fieldwork (see “Methods” section). Table 1 presents summary statistics of the patient population based on these geographic analyses. Though more than two thirds of the population lived further than 5 km from a PHC and 27% lived further than 10km, these populations represented only 40% and 9% of all patient visits respectively. Only a fourth of the population lived within 1h of a PHC. Average annual PHC utilization per capita rates were nearly triple inside the HSS intervention catchment (0.64) than in the rest of the district (RoD, 0.23) for all ages, and more than double for children under 5 years. Utilization rates more than halved for every 5km and every hour of travel from a PHC for every age group considered (Table 1).

**Table 1.** Geographic distribution of populations and PHC outpatient visits in Ifanadiana district, 2014-2017

|  | District | HSS Catchment |  | Distance to PHC |  |  | Time to PHC |  |  |
| --- | --- | --- | --- | --- | --- | --- | --- | --- | --- |
|  |  | Inside | RoD | 0-5 km | 5-10 km | 10-22 km | 0-1 h | 1-2 h | 2-5 h |
| Population (prop.) |  |  |  |  |  |  |  |  |  |
| All Ages | 198,175 | 72,152<br>(0.36) | 126,023<br>(0.64) | 63,811<br>(0.32) | 80,790<br>(0.41) | 53,573<br>(0.27) | 49,269<br>(0.25) | 74,169<br>(0.37) | 74,738<br>(0.38) |
| Under 5 years old | 35,671 | 12,987<br>(0.36) | 22,684<br>(0.64) | 11,486<br>(0.32) | 14,542<br>(0.41) | 9,643<br>(0.27) | 8,868<br>(0.25) | 13,350<br>(0.37) | 13,453<br>(0.38) |
| Over 5 years old | 162,503 | 59,164<br>(0.36) | 103,338<br>(0.64) | 52,325<br>(0.32) | 66,248<br>(0.41) | 43,930<br>(0.27) | 40,400<br>(0.25) | 60,818<br>(0.37) | 61,285<br>(0.38) |
| Total number of patients (prop.) |  |  |  |  |  |  |  |  |  |
| All Ages | 270,747 | 173,497<br>(0.64) | 97,250<br>(0.36) | 163,656<br>(0.6) | 83,667<br>(0.31) | 23,424<br>(0.09) | 145,249<br>(0.54) | 83,154<br>(0.31) | 42,344<br>(0.15) |
| Under 5 years old | 92,533 | 52,240<br>(0.56) | 40,293<br>(0.44) | 52,761<br>(0.57) | 31,308<br>(0.34) | 8,464<br>(0.09) | 45,576<br>(0.49) | 31,356<br>(0.34) | 15,601<br>(0.17) |
| Over 5 years old | 178,214 | 121,257<br>(0.68) | 56,957<br>(0.32) | 110,895<br>(0.62) | 52,359<br>(0.29) | 14,960<br>(0.08) | 99,673<br>(0.56) | 51,798<br>(0.29) | 26,743<br>(0.15) |
| Per capita utilization per year |  |  |  |  |  |  |  |  |  |
| All Ages | 0.39 | 0.64 | 0.23 | 0.74 | 0.29 | 0.12 | 0.84 | 0.32 | 0.16 |
| Under 5 years old | 0.74 | 1.07 | 0.53 | 1.33 | 0.6 | 0.25 | 1.46 | 0.67 | 0.33 |
| Over 5 years old | 0.31 | 0.54 | 0.16 | 0.61 | 0.22 | 0.1 | 0.70 | 0.24 | 0.12 |

**Figure 1.**
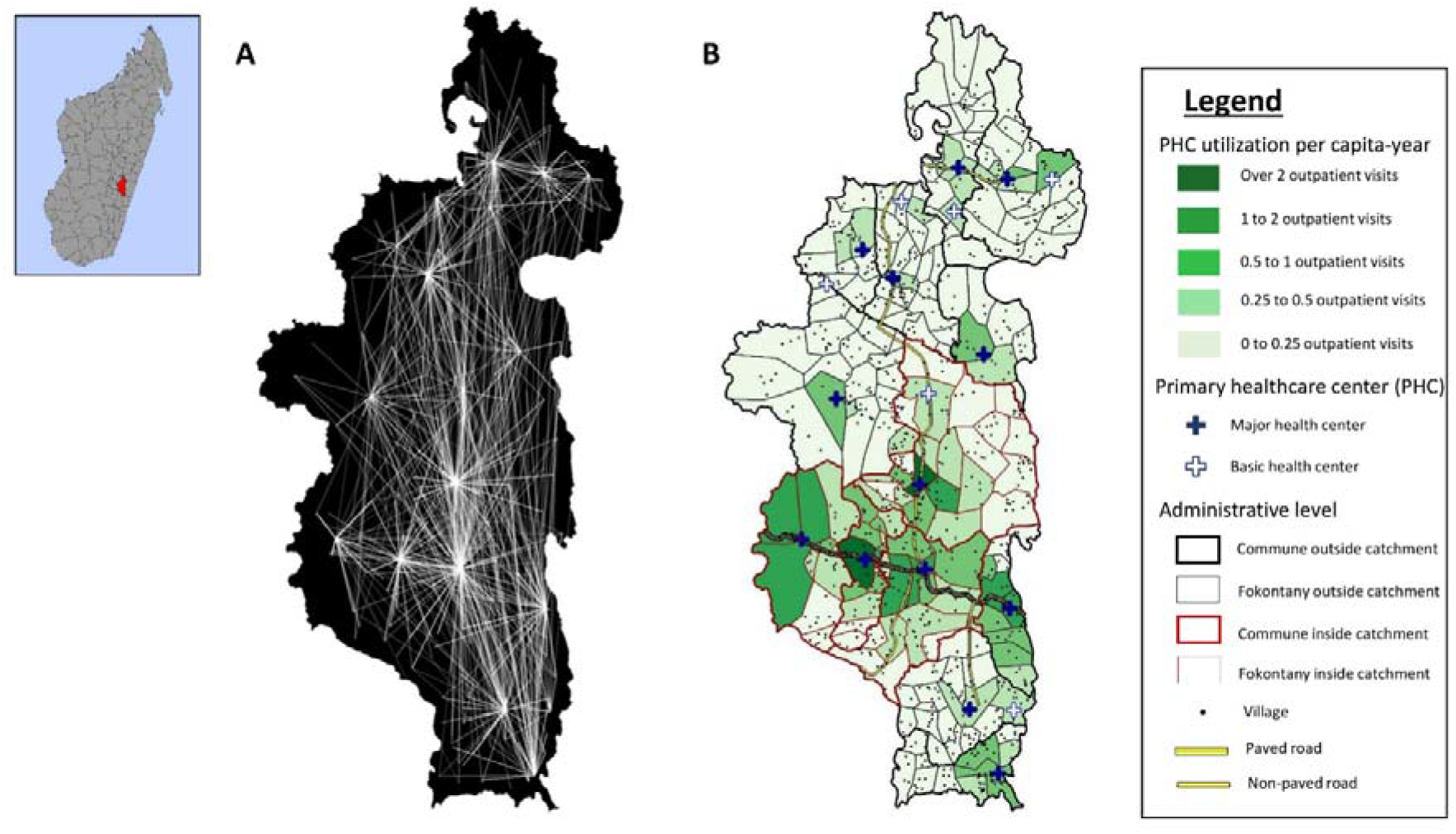
Geographic origin of patients from the 19 PCH in Ifanadiana district, 2014-2017. (A) Origin of all patients attending each of the 19 health centers in Ifanadiana district. For improved visualization, lines between Fokontany and PHC are included only when they represent over 100 patients, with line transparency inversely proportional to the logarithm of the number of visits. These data were aggregated to obtain a total number of per-capita visits per month for each Fokontany (lowest administrative unit, comprising one or several villages). (B) Average number of PHC visits per capita-year for each Fokontany during the study period.

Spatial analyses revealed that utilization rates increased over time in the HSS intervention catchment but declined dramatically as distance increased within the first 5km from a PHC, especially after the system was strengthened at the facility level (Figure 2). The HSS intervention exacerbated the impact of geography on utilization (Figure 2B and 2C), but the ratio in PHC utilization between populations living in close proximity (<2.5km) versus those leaving the furthest (>10km) remained the same at over 10 times higher. Following user fee exemptions, the HSS intervention catchment experienced a substantial increase in utilization, from 1 to nearly 3 visits per capita-year for populations living in close proximity to a PHC, and from 0.25 to about 0.5 visits per capita-year for populations living 5-10 km from a PHC (Figure 2B and 2C).

**Figure 2.**
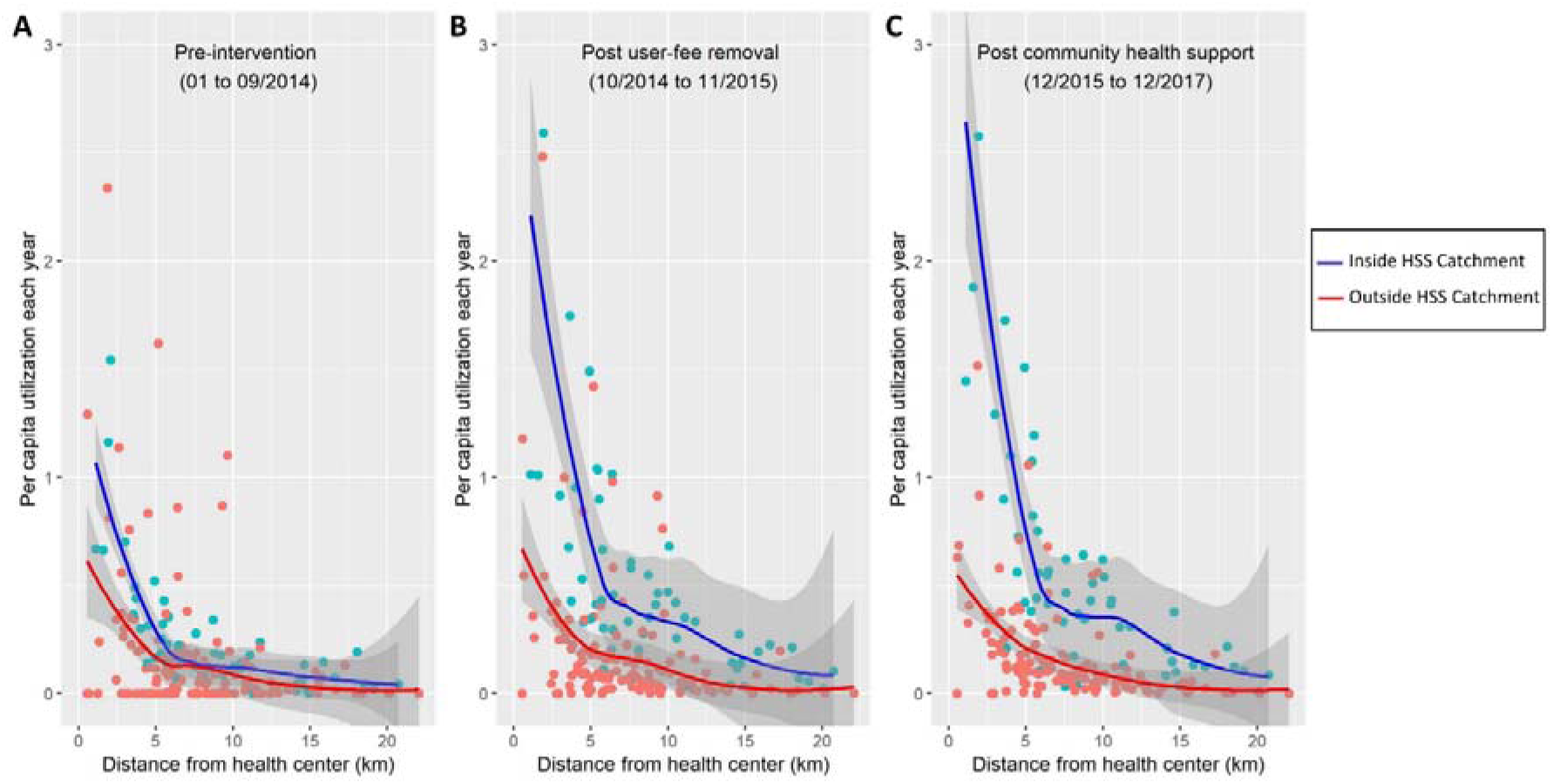
Average PHC per capita utilization by distance of Fokontany to PHC. Colors represent the HSS intervention catchment (blue) and the rest of Ifanadiana district (blue). Each dot represents one of the 195 Fokontany in Ifanadiana, solid lines are the respective non-linear smooth (local regression, LOESS method) and grey shades are the 95% confidence intervals around each smooth. A clear distance decay pattern can be observed, accentuated for the HSS intervention catchment due to a larger increase in utilization near PHC following the HSS intervention (B and C). To improve visualization, one dot from the HSS intervention catchment post intervention (4.5 per capita-year) was removed.

Despite strong seasonal variation in utilization rates, particularly for populations living close to a PHC, trends remained unchanged in the RoD during the study period (Figure 3). Compared to the RoD, HSS activities in the intervention catchment resulted in a shift by 5km in PHC utilization patterns so that populations living 5km further from a strengthened PHC accessed care at rates comparable to those living 5km closer to a facility that did not receive the intervention (Figure 3). We also observed seasonality in the average distance patients travelled to access a PHC during the year (Figure S2). Overall, 50% of outpatient visits seen in the intervention catchment came from localities within 2km of a HCF and over 75% from localities within 4km, but in the months of May through July (dry season) patients came from further away. These seasonal patterns were not observed in the RoD (Figure S2).

**Figure 3.**
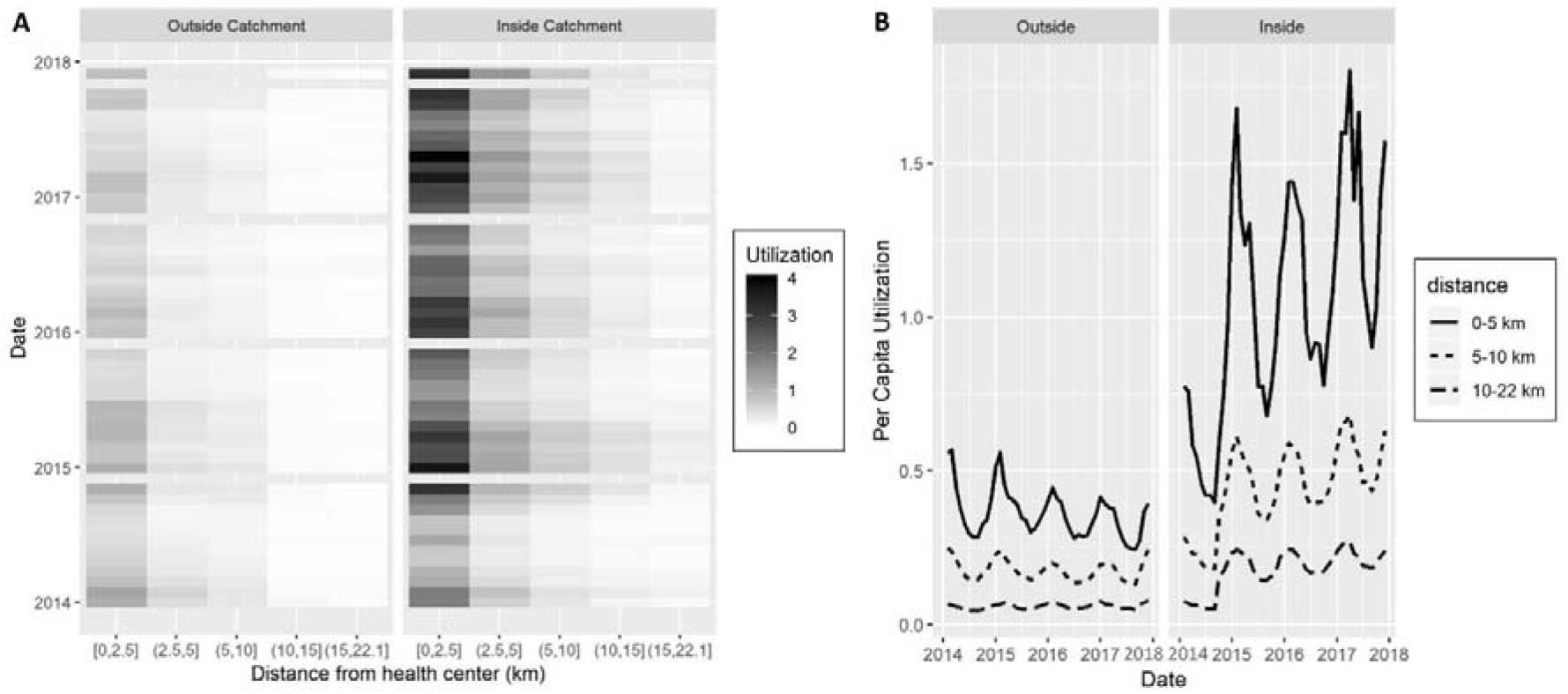
Time-series of PHC per capita utilization by distance of Fokontany to PHC, inside and outside the HSS intervention catchment. (A) Heat map of observed monthly PHC utilization, where grey color scale is proportional to average per capita values at each distance. (B) Model predictions of PHC per capita utilization for Ifanadiana, aggregated by intervention catchment area and distance to PHC. Both graphs show that implementation of HSS activities in the intervention catchment resulted in a shift by 5km of PHC utilization patterns (e.g. those living 5-10km from a PHC in the intervention catchment, have similar utilization rates than those within 5km outside the catchment). Utilization rates were annualized to improve comparability of results.

### Impact of HSS programs on PHC utilization

We analyzed the main factors associated with PHC utilization rates at each Fokontany over time using interrupted time-series analyses with control groups (see “Methods” section). Our models confirmed the exponential decrease in PHC utilization due to geographic distance after accounting for program implementation, health system factors and underlying temporal trends (Table 2, Figure S3). We carried out several models to understand the consistency of associations when including or excluding malaria cases (because of their influence on PHC utilization seasonality), as well as for populations of all ages or only children under 5 years. In every model, a non-linear relationship with distance to PHC best explained utilization patterns (better than travel time or a linear relationship with distance), and this was the most important variable associated with PHC utilization trends (Table S2). After controlling for time trends and baseline differences in health system factors, patterns of geographic utilization of health care services were also highly sensitive to HSS programs implemented in this period, especially the fee-exemption program to increase financial access to PHC, and the community program to address geographic barriers (Table 2). Both programs had a positive impact on PHC utilization rates for all ages (OR=1.09 and OR=1.14 respectively), with a higher relative increase for those populations living further away (OR=1.45 and OR=1.09 respectively, every 10km from a PHC). These results were consistent regardless of the age group considered or whether malaria cases were included in the model (Table 2). Our models accurately explained spatial and temporal utilization patterns at PHC (Figure S4), allowing us to predict dynamics of PHC geographic utilization in the district (S2 Video).

**Table 2.** Multivariate model results (Generalized linear mixed models with random intercept at the PHC closest to the Fokontany of residence)

| Variable | Outpatient visits for all ages | Outpatient visits for children under age 5 | Outpatient visits for all ages, excluding malaria | Outpatient visits for children under age 5, excluding malaria |
| --- | --- | --- | --- | --- |
|  | OR (95% CI) | OR (95% CI) | OR (95% CI) | OR (95% CI) |
| Intercept (Visits per capita-month) | 0.02 (0.011-0.037) | 0.032 (0.019-0.054) | 0.015 (0.008-0.03) | 0.024 (0.013-0.043) |
| **Geographic factors** |  |  |  |  |
| Network distance to PHC (10km, linear)<sup>a</sup> | 0.126 (0.121-0.131) | 0.228 (0.212-0.244) | 0.095 (0.091-0.1) | 0.141 (0.13-0.154) |
| Network distance to PHC (10km, quadratic)<sup>a</sup> | 1.102 (1.076-1.13) | 0.898 (0.862-0.937) | 1.261 (1.226-1.297) | 1.07 (1.018-1.126) |
| **Health system factors** |  |  |  |  |
| Number of health staff | 1.042 (1.037-1.048) | 1.032 (1.023-1.041) | 1.043 (1.037-1.049) | 1.039 (1.029-1.048) |
| Major PHC (vs. basic PHC) | 3.238 (1.5-6.993) | 3.449 (1.764-6.744) | 3.16 (1.421-7.024) | 3.634 (1.762-7.493) |
| HSS catchment (vs. outside) | 0.663 (0.636-0.69) | 0.637 (0.597-0.68) | 0.641 (0.611-0.672) | 0.634 (0.587-0.684) |
| **Impact of HSS programs** |  |  |  |  |
| User fee exemption program | 1.09 (1.063-1.118) | 0.946 (0.908-0.987) | 1.18 (1.145-1.215) | - |
| Interaction with Distance to PHC (10km)<sup>a</sup> | 1.454 (1.433-1.476) | 1.416 (1.385-1.448) | 1.44 (1.415-1.466) | 1.361 (1.328-1.394) |
| Community health program | 1.147 (1.125-1.17) | 1.148 (1.118-1.179) | 1.112 (1.087-1.137) | 1.081 (1.039-1.125) |
| Interaction with Distance to PHC (10km)<sup>a</sup> | 1.095 (1.08-1.111) | - | 1.127 (1.109-1.145) | 1.052 (1.02-1.085) |
| **Underlying trends** |  |  |  |  |
| Linear trend (year) | 0.968 (0.962-0.973) | 0.968 (0.959-0.977) | 0.988 (0.981-0.995) | - |
| Seasonal trend<sup>b</sup> | 1.204 (1.197-1.211) | 1.212 (1.199-1.225) | 1.031 (1.024-1.038) | 1.127 (1.114-1.141) |
| Lagged trend (1-month lag)<sup>c</sup> | 1.473 (1.466-1.48) | 1.304 (1.297-1.311) | 1.65 (1.637-1.663) | 1.363 (1.352-1.375) |

<sup>a</sup> The variable network distance represents tens of kilometers (distance in km\*10<sup>-1</sup>) to facilitate interpretation of coefficients and enable model convergence
<sup>b</sup> Seasonal trend was constructed as $\sin(2\pi(\text{Month}_i + \theta/12))$ , where $\theta$ was the horizontal shift that best fit the data of each model.
<sup>c</sup> Lagged trend transformed into visits per capita-year to allow interpretation of results

Predictions from the model for all ages suggested that in the absence of these programs, only 1% of the population in Ifanadiana district would have per capita PHC utilization of 1 visit or more per year, and 12% would have 0.5 visits or more per year. If these programs were implemented everywhere in the district, nearly one quarter of the population (23%) would have a PHC utilization of at least 1 visit and nearly half (47%) would have at least 0.5 visits per capita-year. Maps in Figure 4 show predictions of the geographic distribution of PHC utilization with and without implementation of HSS programs, revealing substantial gaps in health system coverage for remote populations. PHC utilization remained low for remote populations under a variety of HSS scenarios that included hiring additional health staff at PHC, removing user fees and strengthening community health (Figure S5).

**Figure 4.**
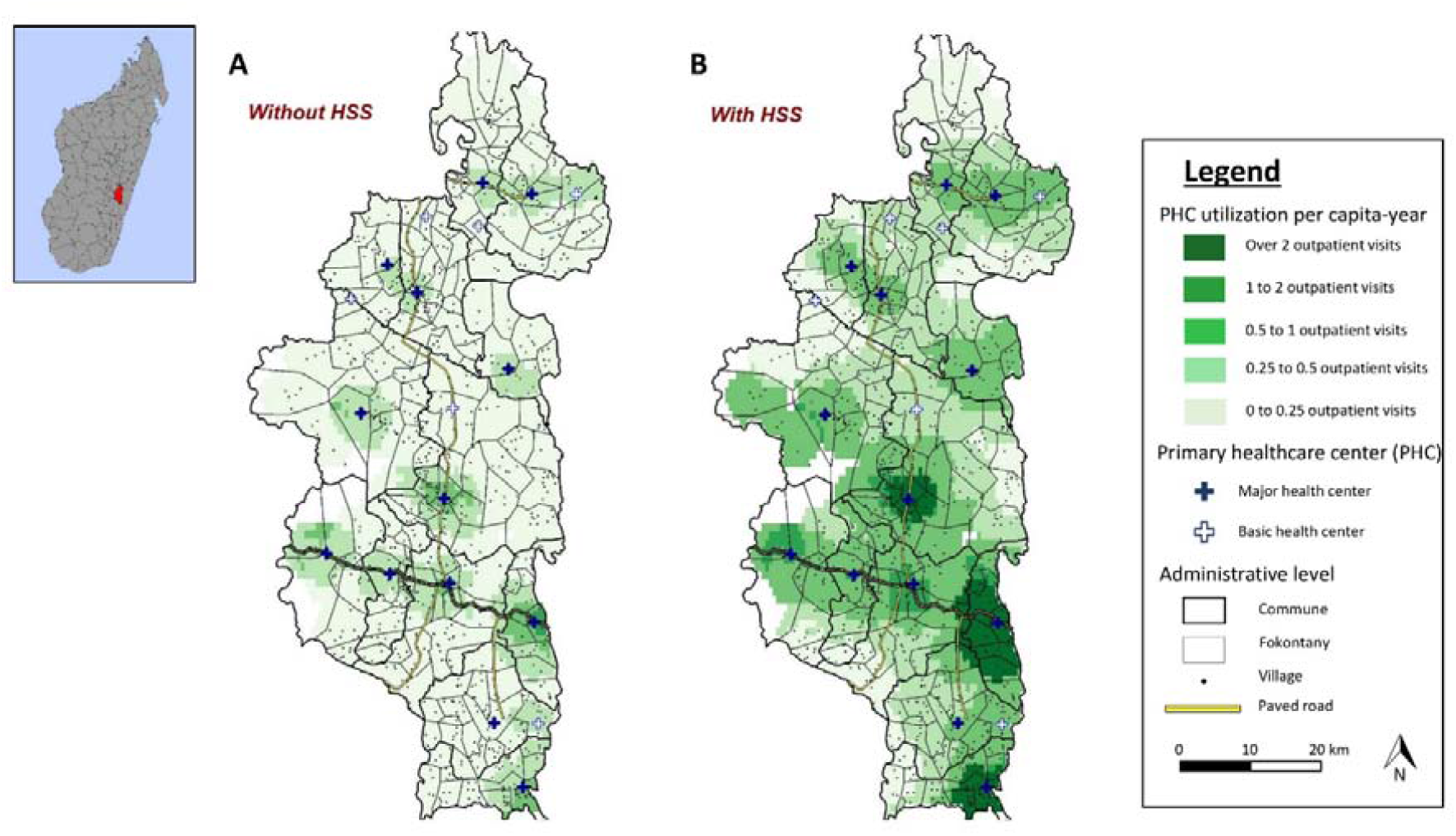
Predictions of geographic distribution of PHC per capita utilization in Ifanadiana district according to scenarios of HSS intervention implementation. Color shades represent predictions of annual PHC per capita visits in (A) a scenario where no HSS activities are implemented, and (B) a scenario where HSS are implemented in the whole district. Maps reveal that despite improvements, even if HSS were implemented across the district, a large proportion of the population would remain with very low levels of realized access to facility-based primary care.

### Utilization for children under 5 years combining PHC and community health data

To reduce geographic barriers to care, community health workers (2 per Fokontany) can manage childhood illnesses such as malaria, diarrhea or pneumonia for children under 5 years of age. To understand how community health consultations can complement facility-based delivery, we collected consultation data from community health sites in four communes of the HSS intervention area from January to December 2017, where the community health program was supported by the HSS intervention. For children under five, utilization of primary care in this period exceeded one visit per child regardless of the distance of the population to a PHC for the 39 of the 43 Fokontany (94% of under 5 population) when combining outpatient visits from both PHC and community health sites (Figure 5A). On average, combined utilization exceeded 2 visits per child per year in all distance groups from a PHC (Figure 5B). Average utilization at community health sites substantially increased at further distances from a PHC: annual utilization was less than 0.5 at 2.5km from a PHC and nearly 2 at Fokontany more than 15km from a PHC. As a result, visits at community health sites accounted for 90% of total primary care visits in Fokontany further than 15km from a PHC, while they accounted for only 10% of total visits at 2.5km or less from a PHC (Figure 5B). Combined utilization of primary care was still lower for children living further away from a PHC, but the effect of distance was much less pronounced due to the exponential increase in community health site utilization at higher distances from PHC (Figure 5A), which essentially compensated for the distance decay.

**Figure 5.**
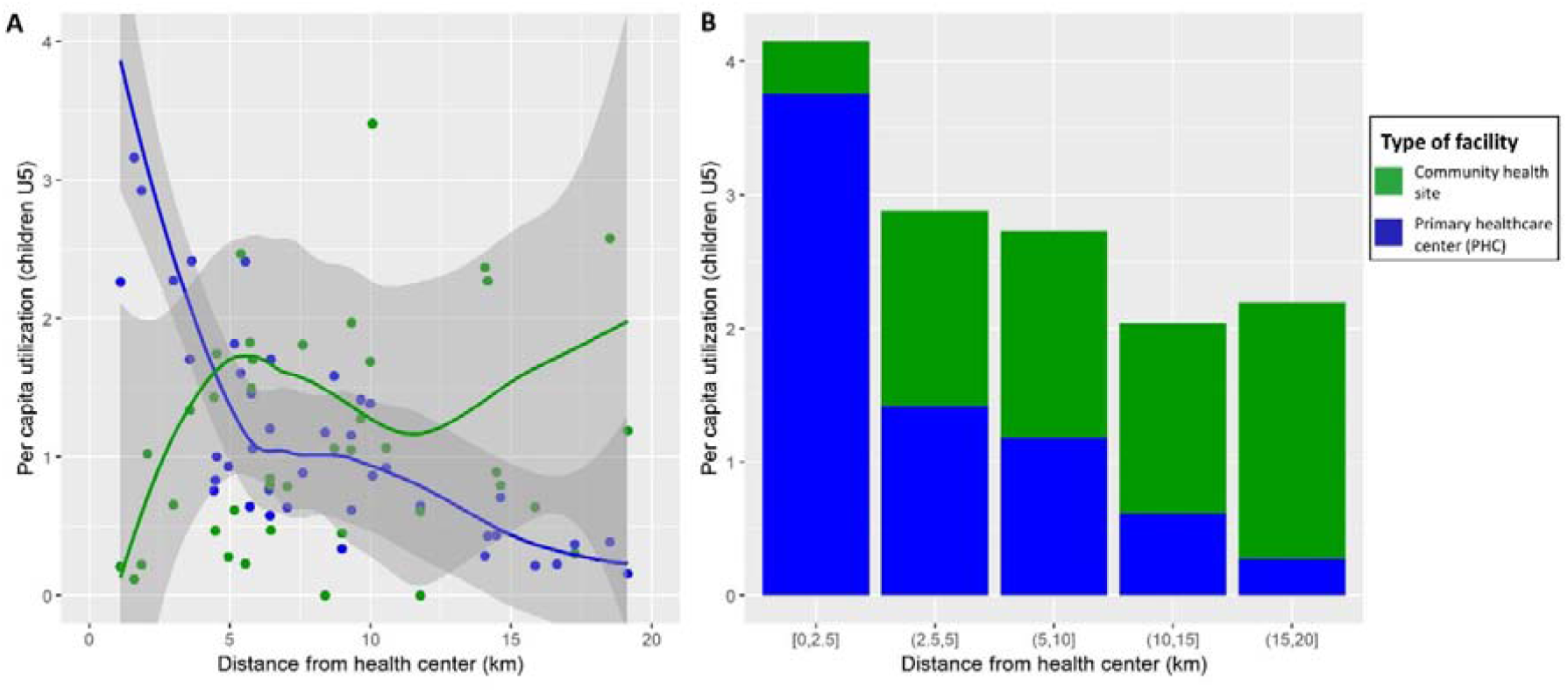
Annual utilization rates of primary care (PHC and community health sites) per capita by children under 5 years in the HSS intervention catchment, 2017. Graphs show per capita utilization at different distances to the nearest PHC, disaggregated by PHC and community health visits. It reveals that utilization at community health sites compensates the distance decay observed for PHC use, with higher community health site utilization at further distance to PHC and reaching over 2 visits per capita-year for all distance groups. To improve visualization, four dots were removed in the left graph (utilization over 4 per capita-year).

## DISCUSSION

A renewed commitment to strengthening primary health care systems and ensuring universal health coverage is essential to meet the health-related Sustainable Development Goals [27], but enormous questions remain on how to do this [20,28–31]. Here, we analysed geographic data from hundreds of thousands of patients in a rural district of Madagascar undergoing health system transformation to understand how facility- and community-based interventions contribute to health system coverage. Our results reveal strong limitations of the reach of facility-based primary care, even when it is free of charge and of improved quality [5,25]. Communities that lived within 5km of a supported PHC exceeded 1 visit per person-year, but the intervention accentuated the distance decay (exponential decrease) in PHC utilization, widening the gap with remote populations. We predict that scaling up PHC interventions alone would only achieve modest increases in geographic coverage. Strengthening community-health can have substantial impacts on the geographic reach of the health system. The effect of geography was greatly reduced for children when combining primary care utilization at PHC and community health sites.

Research on geographic accessibility to care has generally focused on characterizing either potential access (population within a certain distance from a PHC) or realized access (actual utilization at different distances to a PHC) [32,33]. In terms of potential access, a travel time of one or two hours to health services is a typically accepted measure of accessibility to health services [13,34–37]. We estimated distance to PHC using a complete district mapping of over 20,000 km of footpaths and 100,000 houses. We then parametrized travel time with hundreds of hours of fieldwork and remote sensing analyses. This approach allowed us to improve on previous methods in developing countries that use either Euclidean distances, friction surfaces [15,38,39], or self-reported answers in surveys [13,14,40]. We found that the majority of the population in Ifanadiana district (75%) lived more than 1h from primary care at a PHC and over one third (38%) lived further than 2h. This is comparable to the proportion lacking access to secondary care at hospitals in other African settings [13,36,37].

When user fees were removed and HSS activities were in place, we found that utilization rates reached between 1 and 3 consultations per person-year for populations in close proximity to PHC, similar to findings in other studies in Africa [8], and close to utilization rates in many OCDE countries with lower disease burdens [41]. Although distance to PHC is not always associated with lower utilization or worse outcomes [39,42,43], we observed a similar distance decay in utilization as previously described in other settings [11–15,44]. Moreover, we show that this decay can be even more pronounced once interventions aimed at increasing healthcare access have been implemented. We found that the median distance of patients to PHC following the HSS was 2 km, similar to results found in a rural area of Western Kenya [11]. Our results are also consistent with evidence from Burkina Faso and Ghana, where user fee exemptions and HSS strategies achieved greater equity across socio-economic groups but did not overcome geographic barriers [7,18,19,45,46].

Using geographic information from nearly 300,000 primary care visits to PHC, we show that health system data can allow for powerful study of spatio-temporal changes in health care access and for drawing key insights to improve UHC strategies. Previous studies that combined measures of geographic access with health care utilization or service coverage have been restricted to discreet services or conditions such as obstetric care, tuberculosis, malaria, or HIV [14,35,39,42,47,48]. Electronic HMIS data currently available rarely include a low level of geographic disaggregation, so studies typically use information from national surveys or restrict the extraction of HMIS geographic information to particular conditions [7,12,18,45,49–51] or to small samples of patients [15,43]. One of the most precise studies linked over 3,000 pediatric health visits in 7 clinics in Kenya to individual identifiers from a demographic surveillance system [11]. However, a push for electronic data collection to improve health information systems is underway in many developing countries with the scale-up of the open source DHIS2 (District Health Information Software) among other platforms, which can be combined with community-based mobile tools for registering cases, and track patient-level data at different levels of care [52]. The level of granularity and timeliness of data of these e-health platforms will open new possibilities for integration of feedback loops between spatial modelling approaches in local planning and implementation of health strategies to maximize geographic access.

The results from this study have implications for the UHC strategy in Madagascar and elsewhere, suggesting that wider support to community health may be necessary to achieve universal access to primary care. However, there remains debate on how to optimize community health. In Madagascar, as in most countries, the national policy considers community health workers as local volunteers, with minimum requirements of formal education and compensation well below the national minimum wage, based on a social marketing strategy. Community-based diagnosis and treatment is restricted to malaria, pneumonia, and diarrhea for children under five. The burden of disease thus remains unmet for the large majority of the population, even when the health system is fully functioning. New WHO guidelines, not yet fully adopted by most countries, recommends paying community health workers minimum wage, removing use fees, and dedicated supervision [20]. In order to directly address the geographic burden of disease, a greater ability to reach facilities is critical, and professionalized community health workers would need to expand the scope of primary care services across a greater range of clinical cases and demographic characteristics.

## METHODS

### Study site

Ifanadiana is a rural health district of approximately 200,000 people located in the region of Vatovavy-Fitovinany, in Southeastern Madagascar. As per Ministry of Health (MoH) norms, Ifanadiana district has one reference hospital, one main primary care health center (PHC2) for each of its 13 communes (subdivision of a district with ∼15,000 people), six additional basic health centers for its larger communes (PHC1), and one community health site with two community health workers for each of its 195 Fokontany (subdivision of a commune with ∼1,000 population). The integrated HSS intervention carried out by the MoH-PIVOT partnership (summarized in Table S1) began in 2014, is guided by existing MoH policies, covers all six WHO building blocks of HSS, and is implemented across all three levels of care in the district (community, health center and hospital). This intervention is structured through the integration of horizontal improvements in system “readiness”, vertically aligned clinical programs, and information systems. Readiness includes infrastructure and sanitation, staffing and equipment to improve the quality of care; procurement systems; an ambulance network; the removal of user fees and provision of social support to patients; trainings and frequent supervision of health staff. The clinical programs include malnutrition and integrated management of child illness (IMCI) through strengthened community health programs, primary health care centers, and hospital (details can be found in [26,53]). The core activities in the first years (2014-2017) covered approximately one third of the population of Ifanadiana (referred to as “PIVOT catchment”), with some activities such as medical staff recruitments spanning the whole district (Table S1).

In addition to PIVOT’s intervention, the population of Ifanadiana benefited from two other notable programs that covered both the PIVOT catchment and the rest of the district (RoD) in this period. The PAUSENS project, funded by the World Bank and implemented in 2013-2017, provided a basic package of services free of charge in all 13 PHC2 for every woman attending the health center for antenatal, delivery, or postnatal care (first six weeks) and children under age five with any illness [54]. The project also included training, support for child vaccination in remote areas, and some equipment to health centers. The Mikolo project, funded by USAID and implemented in 2012-2017, provided support to a network of 150 community health workers in the remote Fokontany (further than 5km from a health center) of eight communes in Ifanadiana, four of which were in the PIVOT catchment and four in RoD. The project organized annual trainings and periodic supervision, provided some equipment, supplies and an initial stock of medicines to each community health workers. The main difference between PIVOT catchment and RoD (our control group) was the implementation of the PIVOT HSS intervention.

### Health system utilization data

From January 2014 to December 2017, we obtained data from the registries on all individuals attending any PHC for an outpatient consultation in the district. The data were collected via regular visits to each PHC in the district by PIVOT staff every 3-4 months, in agreement with the head of each PHC and the district medical inspector. This allowed for the creation of a patient-level, de-identified digital database. For each patient (new visits; follow-up excluded), information included the age, name of the Fokontany of residence and malaria status. Fokontany are the smallest administrative units, comprised of one or several villages, and are located at varying distances from the nearest PHC (0 to 20km). For the period from January to December 2017, we also collected consultation data from the community health sites supported by PIVOT at this time (four out of five communes in the intervention area, estimated population about 55,000). This information, which was already available at the Fokontany level, was obtained from the monthly report to the district and was verified and corrected by the PIVOT monitoring and evaluation team for data quality.

Population data for each Fokontany was obtained from the MoH. Consistent with MoH estimates, the population of children under five years was set at 18% of the total catchment population. Although official population data is sometimes deemed inaccurate, we previously showed that estimating catchment populations using available data for our district from other recognized sources such as WorldPop [55] did not change the results of per capita utilization rates analyses [5]. Information about key dates of the health system strengthening intervention, especially the beginning of the user-fee exemption program and the community health program for each supported commune, were obtained from internal records within the NGO. Number of health professionals at each PHC per month were obtained from district’s records and Service Availability and Readiness Assessments [25]. Use of MoH data for this study was authorized by the Secretary General of the MoH, by the Medical Inspector of Ifanadiana district, and by Harvard’s Institutional Review board.

### Geographic information system

We gathered geographic information from multiple sources in order to estimate the distance and travel time from each house in Ifanadiana district to the nearest PHC. First, we mapped all footpaths, residential areas, houses and rice fields in the district using very high resolution satellite images available through OpenStreetMap (OSM). For this, we implemented a participatory approach in collaboration with the non-profit organization Humanitarian OpenStreetMap Team (HOT), where the district was divided in tiles of 1km by 1km and a request for mapping them was made publicly available through the HOT website [56]. Mapping was carried out in a two-stage process, where tiles that had been mapped had to be validated by a separate contributor. Most tiles were mapped and validated by a dedicated team hired through the project to ensure data quality and completion within the project deadlines. When mapping was completed, we used the Open Source Routing Machine (OSRM) engine to query our OSM data and accurately estimate the shortest path between each house in the district and the nearest PHC.

Second, to estimate travel speed by foot under different terrain and environmental conditions, we conducted field GPS tracking between September 2018 and April 2019 in a sample of itineraries in Ifanadiana. A total of 168 itineraries by foot amounting to nearly 1,000 km were collected by PIVOT community and research teams, in collaboration with community health workers. For this, we used the android mobile app “OSMAnd” installed in tablets (Samsung Galaxy A10.1), and we recorded every 10 seconds the GPS coordinates, time and altitude.

Third, we built remotely sensed land cover maps combining information from Sentinel-2 satellites and OSM. We integrated land cover maps with the rest of GIS data (climate, elevation, etc.) to statistically model travel speed and estimate terrain characteristics associated with higher or lower speed using a generalized additive mixed model. We finally combined model results, GIS data and the shortest paths estimated by OSRM in order to predict travel time to seek care at the nearest PHC for every house in Ifanadiana. The aggregated distance and travel time for a Fokontany was the average of all houses in the Fokontany. A detailed description of the methods used to estimate distance and travel time to PHC is available in Ihantamalala *et al*. (2020) [57].

### Data analysis of health system data

The impact of the HSS intervention on utilization rates at each Fokontany was modelled using interrupted time-series analyses with control groups [58]. For this, we first aggregated health center patient-level information to estimate per capita utilization rates per month for each Fokontany in Ifanadiana district. We studied the linear and non-linear effect of travel distance and travel time from each Fokontany to the nearest PHC on utilization rates. We assessed the impact of two programs designed to reduce financial barriers (i.e., user-fee exemption) and to reduce geographic barriers (i.e., community program) by assessing the level of change in utilization (immediate impact) associated with each program [58]. We controlled for linear and seasonal trends in utilization rates in the absence of the programs; for baseline differences in HSS-supported PHC and in the type of PHC (PHC1 or PCH2); and for the number of health staff over time in the closest PHC for each Fokontany.

Per capita utilization rates at PHC were modelled for each Fokontany using Binomial regressions in generalized linear mixed models (GLMM), with a random intercept introduced for the closest PHC. All other variables were introduced as fixed effects. Each explanatory variable was studied through univariate analyses, and those with p-values bellow 0.1 were included in multivariate models. Quadratic terms were included to account for the non-linear relationship of per capita utilization with travel distance/time to the PHC. We included interaction terms between the HSS programs and the travel distance/time to the PHC in the multivariate model to test whether these programs had a different effect on remote populations. Model selection was performed through step-wise procedures based on AIC, by selecting the reduced model with the lowest AIC. Model assumptions in the final model were verified, including violations to homogeneity and independence of residuals. We introduced a 1-month utilization lag in the final models to remove temporal autocorrelation in the residuals. To facilitate interpretation of results, we report exponentiated model coefficients, which reflect the ratio of change (odds ratio, OR) in utilization rates associated with each explanatory variable. Several sets of analyses were carried out in order to study PHC utilization separately in the general population, in specific age groups (children under 5), and including or excluding malaria cases from the analyses. Using the final model for the general population, we predicted utilization for every Fokontany in Ifanadiana under a scenario without HSS programs (no user-fee exemption, no community health support, 3 health staff per PHC) and with HSS programs (user-fee exemption, community health support, 7 health staff per PHC). Analyses were performed with R software [59], and R packages “lme4”, “gstat”, “rgdal”, and “ggplot2”.

## Data Availability

Data belongs to the Madagascar Ministry of Public Health. For data requests, please contact

## ACKNOWLEDGEMENTS

We are grateful to everyone who contributed to the participatory mapping of Ifanadiana, especially Jérémy Commins, and Blake Girardot for setting the HOT projects online. We thank the staff of the local Ministry of Health team in Ifanadiana district as well as PIVOT’s monitoring and community teams for their support during data collection. Thanks are due to Benjamin Andriamihaja and Tanjona Andréambeloson for their help at different stages of the project.

## AUTHOR CONTRIBUTIONS

Conceived and designed the experiments: AG, CR, LFC, VH, MHB. Performed the analysis: AG, FAI, CR, MR, VH, MHB. Contributed reagents/materials/analysis tools: AG, CR, VH, MHB. Wrote the paper: AG, FAI, CR, LFC, MR, BR, FHR, KEF, JCA, JR, VH, MHB.

## SUPPLEMENTARY INFORMATION

**S1 Supplementary information**. It contains details for the health system strengthening intervention, univariate model results, as well as six supplementary figures.

**S2 Video of geographic health center utilization over time**. It shows geographic changes in monthly PHC utilization per-capita in Ifanadiana district (in-sample model predictions inside and outside the HSS catchment, annualized). For reference, removal of user fees in the initial HSS intervention catchment (in red) took place in October 2014, and was expanded to one additional commune in October 2017.

